# Risk Stratification for Endometrial Cancer Reveals Independent Contributions of Polygenic Risk and Body Mass Index

**DOI:** 10.1101/2025.02.19.25322538

**Authors:** Xuemin Wang, Laure Dossus, Marc J. Gunter, Emma J. Crosbie, Jue-Sheng Ong, Dylan M. Glubb, Tracy A. O’Mara

## Abstract

**Background:** Obesity is a major risk factor for endometrial cancer, but it is unknown whether it impacts the association between genetic risk and endometrial cancer. We incorporated polygenic risk score and epidemiological risk factors in the prediction of and investigated associations of BMI and polygenic risk score with endometrial cancer risk

**Methods:** We generated polygenic risk score for endometrial cancer in 129,829 unrelated female participants of European ancestry (including 956 incident cases with endometrial cancer) in the UK Biobank and predicted endometrial cancer using endometrial cancer polygenic risk score and established epidemiological risk factors, including BMI. We evaluated the performance of endometrial cancer prediction models by odds ratios and area under the receiver operating characteristic curves (AUCs) to using logistic regression. Individual and joint associations of BMI and polygenic risk score with endometrial cancer were assessed using Cox proportional hazards models.

**Results:** An integrated model incorporating both polygenic risk score and epidemiological risk factors achieved a modest, but statistically significant, improvement in predicting endometrial cancer status compared with the model that included epidemiologic risk factors alone (AUC = 0.74 versus 0.73; *P* = 3.98 × 10^−5^). Obese participants (BMI ≥ 30 kg/m^2^) in the top polygenic risk tertile had the highest endometrial cancer risk. We observed independent effects of genetic risk and BMI on endometrial cancer risk.

**Conclusion:** Integrating polygenic risk score with epidemiological risk factors may offer insights into population stratification for endometrial cancer susceptibility. Higher endometrial cancer polygenic risk is associated with endometrial cancer, irrespective of BMI.

## Background

Endometrial cancer is the most common gynaecological cancer in developed countries, with 420,242 new cases and 97,704 new deaths estimated globally in 2022.^1^ Notably, over the last three decades, the incidence and mortality of endometrial cancer has been increasing worldwide.^2^

Previous studies have used epidemiological risk factors including age, BMI, parity, duration of oral contraceptive use, age at menarche, age at menopause, to predict endometrial cancer status at moderate accuracy, with AUC values varying from 0.61 to 0.77 depending on the risk factors included and specific study populations.^3–6^ Polygenic risk score (PRS) approaches, the cumulative dosage effects of multiple genetic variants identified by genome-wide association studies (GWAS), hold promise for disease stratification.^7^ Incorporating polygenic risk scores into epidemiological models have achieved marginal improvement in the prediction of endometrial cancer development.^3,5,6^ However, those studies have only included genome-wide significant or sub-genome-wide significant variants into the construction of the endometrial cancer PRS.

Obesity, typically measured by BMI, is the strongest known modifiable risk factor for endometrial cancer.^8^ With the rising global prevalence of obesity,^9^ the incidence of endometrial cancer is further expected to increase. In addition to obesity and genetic variation, factors such as age, ages at menopause and menarche, parity and endogenous sex hormone levels also affect endometrial cancer risk.^8,10–19^ Despite the impact of all these factors, the aetiology of endometrial cancer is not fully understood and there is a significant gap in research examining the integration of risk factors for endometrial cancer prediction. Indeed, the identification of individuals at an elevated risk of endometrial cancer, irrespective of strong risk factors such as obesity, may be crucial for early detection and facilitate the implementation of targeted screening and preventive strategies. This study uniquely evaluates the combined use of PRS and established risk factors in the prediction of endometrial cancer and explores the individual and joint effects of BMI and genetic susceptibility on endometrial cancer risk in the UK Biobank.

## Methods

### Study Population

A flowchart outlining the study design and population is presented in **Figure 1**. This cohort study was based on data from the UK Biobank, which is a prospective cohort with extensive phenotypic and genotypic data for over 500,000 UK participants aged 40-70 at enrolment. Details of the UK Biobank can be found in Bycroft et al. (2018).^20^ Briefly, participants were genotyped using either UK BiLEVE Axiom Array (807,411 genetic variants) or UK Biobank Axiom Array (825,927 genetic variants). Genetic variants were imputed using the 1000 Genomes phase 3, the UK10K, and the Haplotype Reference Consortium datasets as the imputation reference panels, which resulted in 93,095,623 autosomal single nucleotide polymorphisms (SNPs), short indels and large structural variants and 3,963,705 variants on the X chromosome.

**Figure 1.**
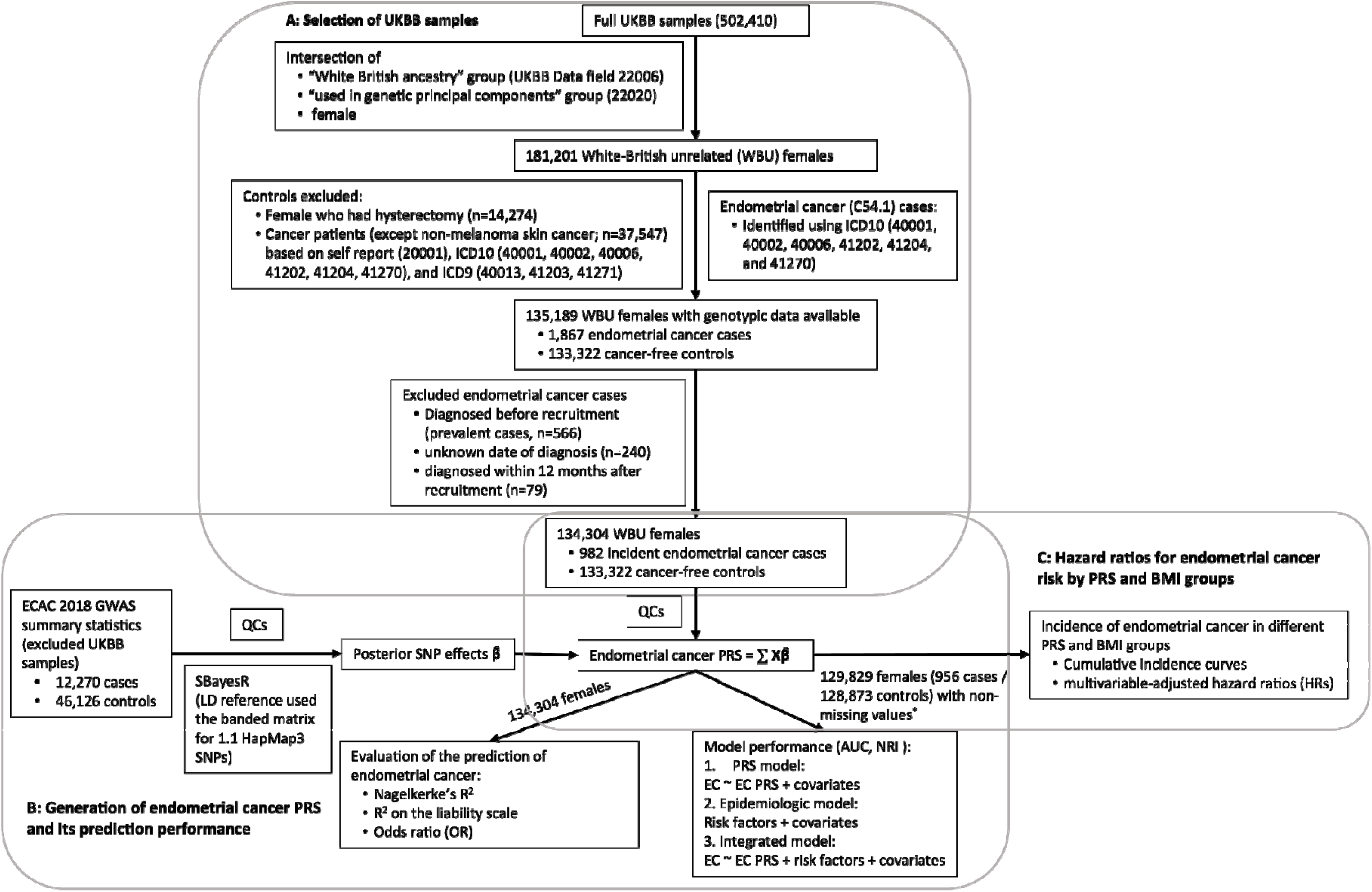
Flowchart for the selection of study participants in the UK Biobank and directed acyclic graph of the study design. (A) Flowchart of inclusion and exclusion criteria. (B) Flowchart of the generation of endometrial cancer polygenic risk scores (PRSs) and evaluation of its prediction of endometrial cancer status. (C) The directed acyclic graph for assessment of endometrial cancer incidence in different BMI and endometrial cancer PRS groups. QCs included filtered out SNPs with MAF < 0.01, multi-allelic variants, missing genotype rate in > 10% of samples, departing from Hardy-Weinberg Equilibrium (*P* < × 10^−6^), and low imputation quality (< 0.4). *4,475 females (26 cases and 4,449 non-cases) within missing values of BMI, age at menarche, number of live births, or ever taken OC pill were excluded.

### Selection of Endometrial Cancer Cases and Cancer-free Participants

Following the exclusion of withdrawn participants, a set of 181,201 unrelated females of European ancestry was defined as the intersection of the “White British ancestry” group (UK Biobank Data Field 22006) and the “used in genetic principal components” group (UK Biobank Data Field 22020) created by Bycroft et al. (2018),^20^ the latter comprising high quality samples filtered to avoid closely related samples (**Figure 1A**).

A total of 1,867 endometrial cancer cases were identified among the 181,201 unrelated females of White British ancestry using the International Classification of Disease 10 (ICD10) subcategory C54.1 (Malignant neoplasm of corpus uteri, Endometrium) in UK Biobank Data Fields 40001 (Underlying (primary) cause of death), 40002 (Contributory (secondary) causes of death), 40006 (Type of cancer), 41202 (Diagnoses – main), 41204 (Diagnoses – secondary), and 41270 (Diagnoses). Diagnosis date was defined as the time of the first endometrial cancer record. From the unrelated females of European ancestry, 133,322 females who had an intact uterus (no hysterectomy) and no prior cancer diagnosis (except non-melanoma skin) were included as controls. Prior cancers were identified based on ICD9 (40013, 41203, and 41271), ICD10 (40001, 40002, 40006, 41202, 41204, and 41270), and self-reported cancers (20001). We restricted analyses to 982 incident endometrial cancer cases. This included the removal of 566 prevalent cases who were diagnosed before recruitment, 240 cases without known date of diagnosis, and 79 cases who were diagnosed within 12 months to mitigate issues related to delayed diagnosis or delayed linkage to cancer registry.

To evaluate endometrial cancer risk prediction models (**Figure 1B & C**), we additionally excluded 4,475 females (26 cases and 4,449 non-cases) with missing values of BMI, age at menarche, number of live births, or ever taken oral contraceptive (OC) pill, resulting in 956 cases and 128,873 cancer-free cohort participants.

### Generation of endometrial cancer PRS

GWAS summary statistics on endometrial cancer risk were sourced from the latest Endometrial Cancer Association Consortium (ECAC) GWAS analysis (12,906 cases and 108,979 controls), which included 636 endometrial cancer cases and 62,853 cancer-free female controls from the UK Biobank.^13^ To avoid potential bias due to sample overlap between the GWAS dataset and the PRS validation dataset, the ECAC GWAS summary statistics were derived again by excluding UK Biobank samples, resulting in 12,270 endometrial cancer cases and 46,126 controls (**Figure 1B**).^21^

All GWAS variants were directly genotyped or well imputed (imputation score > 0.4) and had a minor allele frequency (MAF) > 1%. Posterior effect sizes for genome-wide variants were derived using the SBayesR^22^ method implemented in genome-wide complex trait Bayesian (GCTB) software.^23^ The linkage disequilibrium (LD) matrix was computed based on 1.1 million HapMap 3 variants using a banded matrix with a window size of 3 cM per SNP in a random sample of 50,000 unrelated UK Biobank samples (https://cnsgenomics.com/software/gctb/#LDmatrices).

For UK Biobank individual-level genotypic data, standard GWAS quality controls were conducted to select genetic variants and samples for endometrial cancer PRS generation following the guidelines outlined by Choi et al. (2020)^24^ (see https://choishingwan.github.io/PRS-Tutorial/). Briefly, genetic variants with a MAF < 0.01, a missing genotype rate exceeding 1%, or departing from Hardy-Weinberg Equilibrium (P < 1 × 10^−10^) were excluded, leaving 8,947,018 variants available for potential construction of the endometrial cancer risk PRS. All samples had no more than 10% missing genotypes. Per-individual PRS was calculated as the genome-wide sum of the per-variant posterior effect size multiplied by allele dosage using PLINK (https://www.cog-genomics.org/plink/).^25^ PRS values for individual females were centered to zero.

### Selection of established endometrial cancer risk factors

Drawing on insights from epidemiological and Mendelian randomization studies of endometrial cancer,^11,18,26,27^ we selected and incorporated seven key risk factors into our prediction models for endometrial cancer status (case or non-case). The selected risk factors were: measured BMI (UK Biobank data field 21001), reported age at menarche (2714), age at natural menopause (3581), number of live births (2734), ever taken OC pill (2784), and levels of SHBG (30830) and testosterone (30850) during the initial assessment visit. The proportion of participants with missing information for variables was very low for BMI, age at menarche, number of live births and ever taken OC pill (missingness range 0.3% - 2.9%). The proportion of missing was greater for SHBG (14.3%) and testosterone levels (19.5%) likely due to issues associated with biospecimen processing, including sample quantity and quality failures. Age at menopause was missing for 3.4% of participants and not available for 42.6% of participants because they had not reached menopause.

In order to generate risk models that would be useful pre-menopausal, we generated a polygenic score (PGS) for age at menopause using the GWAS summary statistics provided by Ruth et al. (2021)^17^ and the same procedure as for the derivation for endometrial cancer PRS described above. We used age at menopause PGS in endometrial cancer risk prediction and in the analysis of associations of endometrial cancer with BMI and PRS. We similarly leveraged PGS methods for SHBG and testosterone levels. The use of PGS for these factors in risk prediction models will allow for replication in other cohorts that typically do not have these measured. PGS for levels of SHBG and testosterone were generated using the female-stratified GWAS summary statistics provided by Ruth et al. (2020)^27^ and the same procedure as for the derivation for endometrial cancer PRS. The resulting PGS values for SHBG and testosterone instead of their measured values were included as covariates to assess the performance of our prediction models and in the analysis of associations of endometrial cancer with BMI and PRS.

Age at initial assessment (UK Biobank data field 21003), and the top 10 genetic principal components (PCs) were included in all prediction models as covariates. To estimate genetic PCs, a genetic relationship matrix among individuals was created using HapMap3 variants and the GCTA-GREML method.^28^ The genetic relationship matrix was used to derive genetic PCs using the GCTA software (version 1.94.1).^29^

### Evaluation of PRS model performance and established risk factors

We assessed the predictive performance of the endometrial cancer PRS model in the UK Biobank using logistic regression to calculate the Nagelkerke’s R^2^ and variance on the liability scale explained by PRS as described previously.^30^ Participants were divided into percentiles based on their endometrial cancer PRS distribution, and their estimated odds ratios (ORs) for endometrial cancer risk were calculated using the middle two deciles (40%-60%) as the reference group. AUCs were reported for the endometrial cancer PRS and the following: 1) each of the seven established endometrial cancer risk factors; 2) the epidemiologic model comprising all seven established endometrial cancer risk factors; and 3) an integrated model combining the epidemiological model with the endometrial cancer PRS. All analyses were adjusted for age at initial visit and the top 10 genetic PCs. We used stratified bootstrap with 2000 replicates to compute their corresponding 95% confidence intervals (CIs). We calculated the net reclassification index (NRI) to quantify the improvement in the reclassification of endometrial cancer cases and non-cases by the integrated model as compared to the traditional epidemiological model (i.e. without PRS).

### BMI and PRS associations with endometrial cancer

UK Biobank incident endometrial cancer cases and non-cases were categorised by BMI (BMI < 25kg/m², 25kg/m² ≤ BMI < 30kg/m², and BMI ≥ 30kg/m²) or endometrial cancer PRS tertiles from non-cases. We evaluated associations of BMI and endometrial cancer PRS with endometrial cancer status using Cox proportional hazard models. We tested the proportional hazards assumption for covariates included in a model fit by testing for independence between the scaled Schoenfeld residuals and time. P-values for trend were estimated using endometrial cancer PRS and BMI as continuous variables. The multivariable models were adjusted with each other for BMI groups and endometrial cancer PRS tertiles, and additionally accounted for age at menarche, number of live births (as a categorical factor), ever taken oral contraceptive pill, PGS for age at menopause, PGS for SHBG levels, PGS for testosterone levels, age at initial assessment, and the top 10 genetic PCs. We additionally performed a sensitivity analysis where multivariable models were adjusted with each other for BMI and endometrial cancer PRS as continuous variables. Follow-up time was calculated from the baseline date (date when attended assessment centre during their initial visit) to the date of endometrial cancer diagnosis or death (whichever occurred first). Cumulative incidence rate of endometrial cancer during the follow up period were generated for the different BMI and endometrial cancer PRS groups. We tested the interactions between BMI and endometrial cancer PRS by adding an interaction term in the multivariable model.

We further explored the joint associations of BMI groups and PRS tertiles with endometrial cancer using the Cox proportional model accounting for all covariates mentioned above. A nine-group comprehensive variable was thus formed, and the first PRS tertile and a normal weight (BMI < 25kg/m^2^) group was considered as the reference group.

All analyses were performed using R software version 4.2.2 (https://www.R-project.org). Distribution of baseline characteristics was assessed by descriptive statistics and compared between case and non-case groups using χ^2^ tests for categorical variables and t-tests for continuous variables. Cox proportional hazards ratio models were conducted using the survival (version 3.5-5) and cumulative incidence rate plots were generated using the survminer (version 0.4.9) R packages. NRI calculations were performed using the PredictABEL R package (version 1.2-4).^31^ AUC were assessed using the pROC (version 1.18.0) R package.^32^ Statistical tests were two-sided with a *P* < 0.05 considered statistically significant. Data analysis was conducted from February 22 to Sept 21, 2023.

## Results

### Baseline Characteristics of Study Population

This analysis comprised 134,304 unrelated female participants of the European ancestry, including 982 cases of endometrial cancer and 133,322 cancer-free cohort participants (**Figure 1A**). Consistent with established knowledge, endometrial cancer cases, in comparison to non-cases, exhibited older age at initial assessment, earlier age at menarche, later age at menopause, lower SHBG levels, higher testosterone levels, and a lower frequency of oral contraceptive pill use (**Table 1**). While the mean number of live births did not significantly differ between cases and controls, a higher proportion of cases were nulliparous compared to non-cases (22.6% vs. 18.9%). As anticipated, we observed a higher prevalence of obesity (BMI ≥ 30 kg/m^2^) among participants with endometrial cancer compared to non-cases (*P* < 2.2 × 10^−16^; **Table 1**).

**Table 1.** Baseline characteristics of participants in the UK Biobank.

| Baseline characteristics | Overall | Cases | Controls | P-value |
| --- | --- | --- | --- | --- |
| Number of female participants | 134304 | 982 (0.7%) | 133322 (99.3%) |  |
| Age at initial assessment, years (SD) | 55.7 (8.0) | 59.5 (6.6) | 55.7 (8.0) | $< 2.2 \times 10^{-16}$ |
| BMI (SD), continuous | 26.9 (5.1) | 30.5 (6.9) | 26.9 (5.1) | $< 2.2 \times 10^{-16}$ |
| BMI categories<sup>a</sup> |  |  |  |  |
| BMI < 25kg/m<sup>2</sup> | 55090 (41.0%) | 210 (21.4%) | 54880 (41.2%) | $< 2.2 \times 10^{-16}$ |
| 25kg/m<sup>2</sup> ≤ BMI < 30kg/m<sup>2</sup> | 48703 (36.3%) | 340 (34.6%) | 48363 (36.3%) |  |
| BMI ≥ 30kg/m<sup>2</sup> | 30128 (22.4%) | 427 (43.5%) | 29701 (22.3%) |  |
| Missing (%) | 383 (0.3%) | 5 (0.5%) | 378 (0.3%) |  |
| Age at menarche, years (SD) | 13.0 (1.6) | 12.7 (1.6) | 13.0 (1.6) | $< 9.5 \times 10^{-10}$ |
| Number of females who reported their age at menarche (%) | 130449 (97.1%) | 961 (97.9%) | 129488 (97.1%) |  |
| Missing (%) | 3855 (2.9%) | 21 (2.1%) | 3834 (2.9%) |  |
| Age at menopause, years (SD) | 50.4 (4.4) | 51.7 (4.3) | 50.4 (4.4) | $< 2.2 \times 10^{-16}$ |
| Number of post-menopausal females (%) | 72459 (54.0%) | 754 (76.8%) | 71705 (53.8%) |  |
| Number of pre-menopausal females (%) | 57218 (42.6%) | 184 (18.7%) | 57034 (42.8%) |  |
| Missing (%) | 4627 (3.4%) | 44 (4.5%) | 4583 (3.4%) |  |
| Age at menopause PGS<sup>b</sup> (SD) | 0 (1.43) | 0.17 (1.44) | 0 (1.43) | $2.1 \times 10^{-4}$ |
| SHBG levels (nmol/L; SD) | 62.5 (30.7) | 50.6 (26.0) | 62.6 (30.7) | $< 2.2 \times 10^{-16}$ |
| Number of females that had detectable SHBG levels | 115035 (85.7%) | 854 (87.0%) | 114181 (85.6%) |  |
| Missing (%) | 19269 (14.3%) | 128 (13.0%) | 19141 (14.4%) |  |
| SHBG PGS<sup>b</sup> (SD) | 0 (0.22) | -0.05 (0.23) | 0 (0.22) | $8.2 \times 10^{-11}$ |
| Testosterone levels (nmol/L; SD) | 1.1 (0.6) | 1.2 (0.6) | 1.1 (0.6) | $5.4 \times 10^{-7}$ |
| Number of females that had detectable testosterone levels | 108095 (80.5%) | 820 (83.5%) | 107275 (80.5%) |  |
| Missing (%) | 26209 (19.5%) | 162 (16.5%) | 26047 (19.5%) |  |
| Testosterone PGS<sup>b</sup> (SD) | 0 (0.34) | 0.05 (0.36) | 0 (0.34) | $2.8 \times 10^{-6}$ |
| Number of live births (SD), count | 1.8 (1.2) | 1.7 (1.2) | 1.8 (1.2) | 0.2 |
| Number of live births categories<sup>a</sup> |  |  |  |  |
| 0 (%) | 25428 (18.9%) | 222 (22.6%) | 25206 (18.9%) | $1.1 \times 10^{-2}$ |
| 1 (%) | 17798 (13.3%) | 118 (12.0%) | 17680 (13.3%) |  |
| > 1 (%) | 90999 (67.8%) | 642 (65.4%) | 90357 (67.8%) |  |
| Missing (%) | 79 (0.1%) | 0 | 79 (0.1%) |  |
| Ever taken oral contraceptive pill<sup>a</sup> |  |  |  |  |
| Yes (%) | 112428 (83.7%) | 712 (72.5%) | 111716 (83.8%) | $< 2.2 \times 10^{-16}$ |
| No (%) | 21645 (16.1%) | 270 (27.5%) | 21375 (16.0%) |  |
| Missing (%) | 231 (0.2%) | 0 | 231 (0.2%) |  |

Abbreviations - SD: standard deviation; BMI: body mass index; PGS: polygenic score; SHBG: sex hormone binding globulin
<sup>a</sup>Percentages may not add up to 100% due to rounding;
<sup>b</sup>PGS (polygenic scores) of age at menopause, SHBG, and testosterone were shifted to mean zero

### Integration of PRS and established risk factors for the prediction of endometrial cancer

The endometrial cancer PRS was strongly associated with endometrial cancer risk amongst White-British unrelated participants (P value < 2.2 × 10^−16^) (**Figure 2A**). The Nagelkerke’s R^2^, a pseudo-R^2^ statistic measuring proportion of variance explained by endometrial cancer PRS, was 0.9% and variance on the liability-scale explained by endometrial cancer PRS was 2.2%. Compared to the middle quintile of participants (40-60%), participants in the top 10% and top 1% of the PRS distribution had a 1.98-fold (*P* = 3.03 × 10^−9^) and 3.06-fold (*P* = 7.10 × 10^−07^) increased risk of developing endometrial cancer respectively (**eTable 1**). The PRS model predicted endometrial cancer status at an AUC of 0.67 (95% CI, 0.65-0.69). While this was slightly higher than for six of the established endometrial cancer risk factors (AUC range 0.65-0.66), it was lower than the prediction accuracy for BMI (AUC = 0.71; 95% CI 0.70-0.73) (**Figure 2B; eTable 2**). The epidemiological model that included all seven established risk factors could predict endometrial cancer status with an AUC of 0.73 (95% CI 0.71-0.74); this accuracy was improved by 1% on the addition of the endometrial cancer PRS (i.e. the integrated model, AUC = 0.74; 95% CI 0.72-0.75; *P* = 3.98 × 10^−5^). The continuous net reclassification improvement for endometrial cancer prediction was 0.25 (95% CI 0.19 - 0.31; P < 1 × 10^−4^) for the integrated model compared with the epidemiological model.

**Figure 2.**
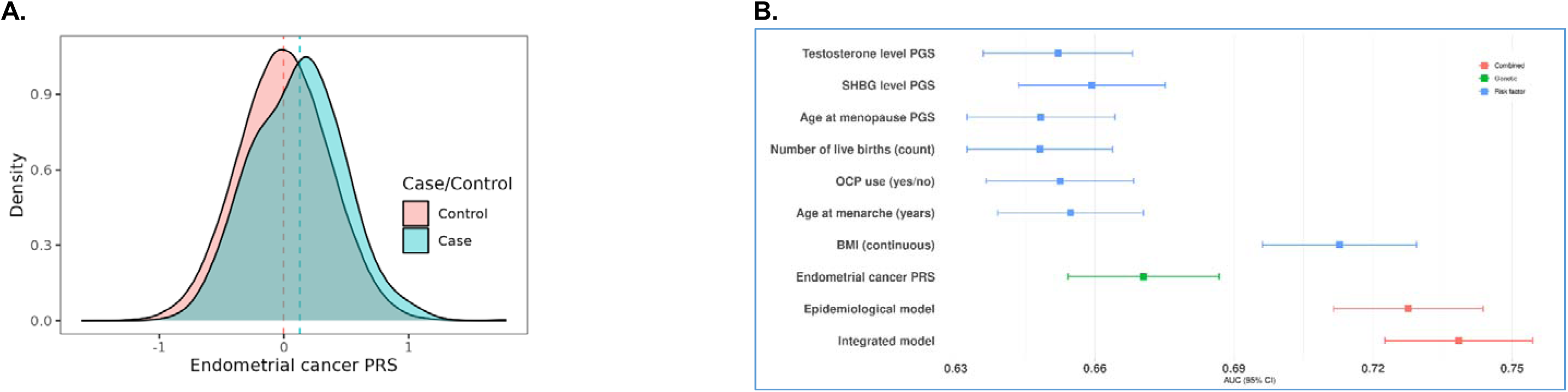
Prediction performance of endometrial cancer polygenic risk score (PRS) in the UK Biobank. (A) The distribution of endometrial cancer PRS by White British unrelated female cases and controls in the UK Biobank. (B) forest plots of AUC of prediction models included endometrial cancer PRS (EC PRS; in green), each individual risk factors (in blue), all risk factors (epidemiological model; in red), and both EC PRS and risk factors (integrated model; in red). Each model was adjusted for age at initial assessment and the top 10 genetic principal components.

### Associations of BMI and PRS with endometrial cancer risk

The proportional hazards regression model assumes that the ratio of hazards between two groups is constant over time. There was no evidence to support violation of the proportional hazard assumption in the current analysis (global test *P* = 0.08). Cumulative incidence curves indicated that only participants in the top tertile of the PRS distribution displayed increased risk compared to the bottom and middle tertiles; whereas higher cumulative incidence rate was associated with both the overweight (25 Kg/m^2^ ≤ BMI < 30 Kg/m^2^) and obese (BMI ≥ 30 Kg/m^2^) groups (**Figure 3A**). BMI and PRS were independently associated with endometrial cancer risk (**Figure 3B**). In the model mutually adjusted for BMI groups and PRS tertiles, compared with the bottom PRS tertile, the top PRS tertile displayed an increased risk (1.71-fold) for endometrial cancer (95% CI, 1.45-2.00; *P* = 3.7 × 10^−11^). For BMI, overweight and obese groups presented a 1.56-fold (95% CI, 1.31-1.86; *P* = 7.2 × 10^−7^) and a 3.03-fold (95% CI, 2.55-3.59; *P* = 4.7 × 10^−37^), increased risk of endometrial cancer, respectively, compared with normal BMI group (**Figure 3B**).

**Figure 3.**
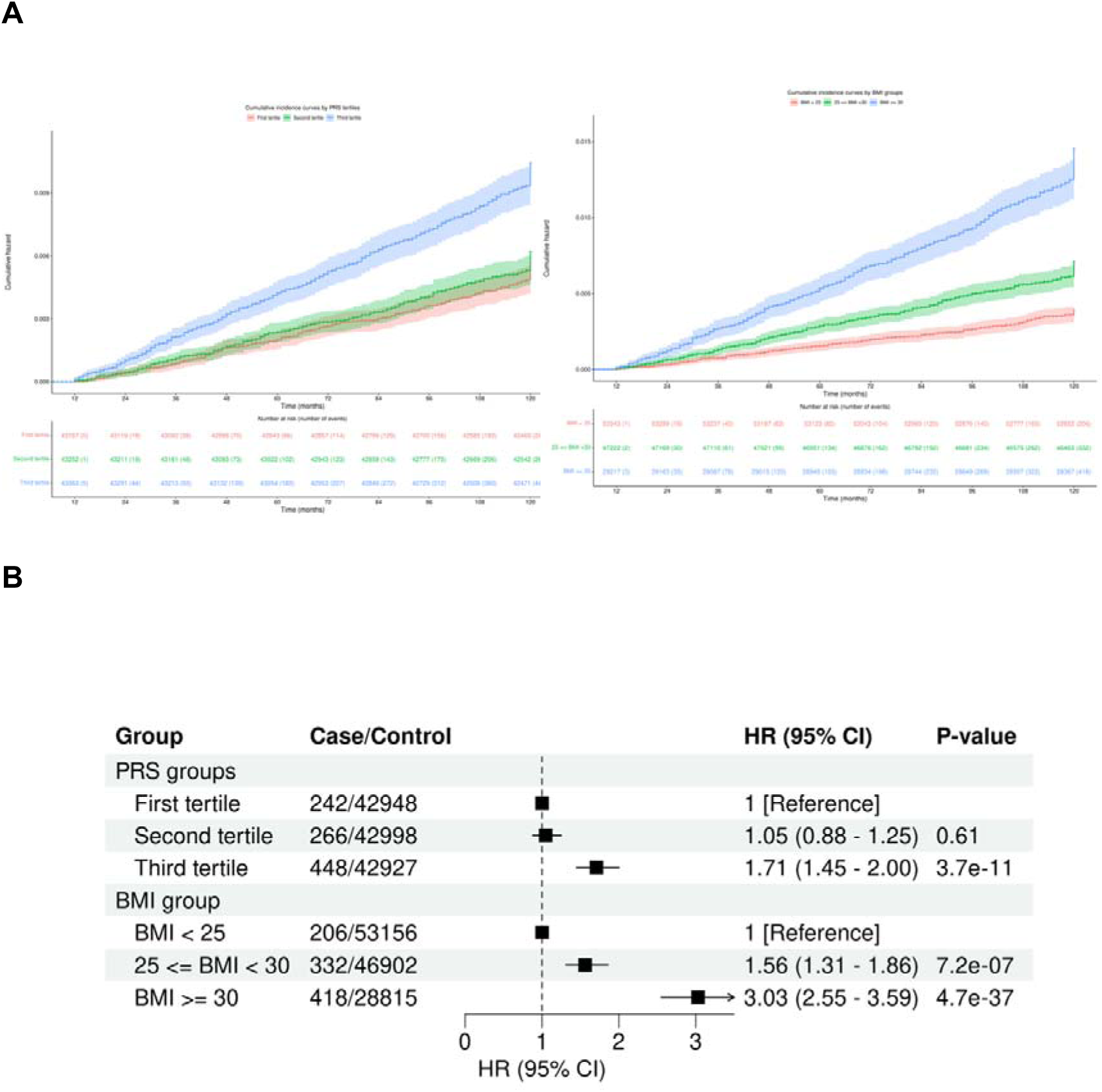
Cumulative incidence curves and multivariable-adjusted effects of endometrial cancer polygenic risk score (PRS) and BMI on endometrial cancer risk. (A) Cumulative incidence curves were drawn for PRS tertiles and BMI groups in the UK Biobank. The unit of follow-up time was months, and the start point was defined as the 12^th^ month (1 year after recruitment). (B) Multivariable models were adjusted for either PRS or BMI group and additionally adjusted for age at menarche, number of live births, ever taken oral contraceptive pill, and PGS values of age at menopause and levels of SHBG and testosterone, as well as age at initial assessment and the top 10 genetic principal components.

When endometrial cancer PRS and BMI were treated as continuous variables, an increase of one standard deviation of endometrial cancer PRS was associated with 2.13-fold risk of endometrial cancer (95% CI, 1.79-2.54; *P* = 2.52 × 10^−17^), while an increase of BMI by 1 kg/m^2^ was associated with a 1.09-fold risk (95% CI, 1.08-1.11; *P* = 2.52 × 10^−80^). There was no evidence of interaction between BMI and endometrial cancer PRS (*P* for interaction = 0.39). Upon stratification by BMI group, we observed that only those with the greatest polygenic load (i.e. the top PRS tertile) in each group had an increased risk of endometrial cancer risk. Participants in the top PRS tertile and the highest BMI group exhibited the greatest risk (HR = 4.94; 95% CI, 3.65-6.68; *P* = 7.67 × 10^−25^; **Figure 4**). Even for participants with a normal BMI, those in the top PRS tertile had a 2.01-fold increased risk (95% CI, 1.45-2.78; *P* = 2.50 × 10^−5^), compared with the bottom PRS tertile. Increased risks for the top PRS tertile persisted after additionally adjusting for continuous BMI (**eFigure1**).

**Figure 4.**
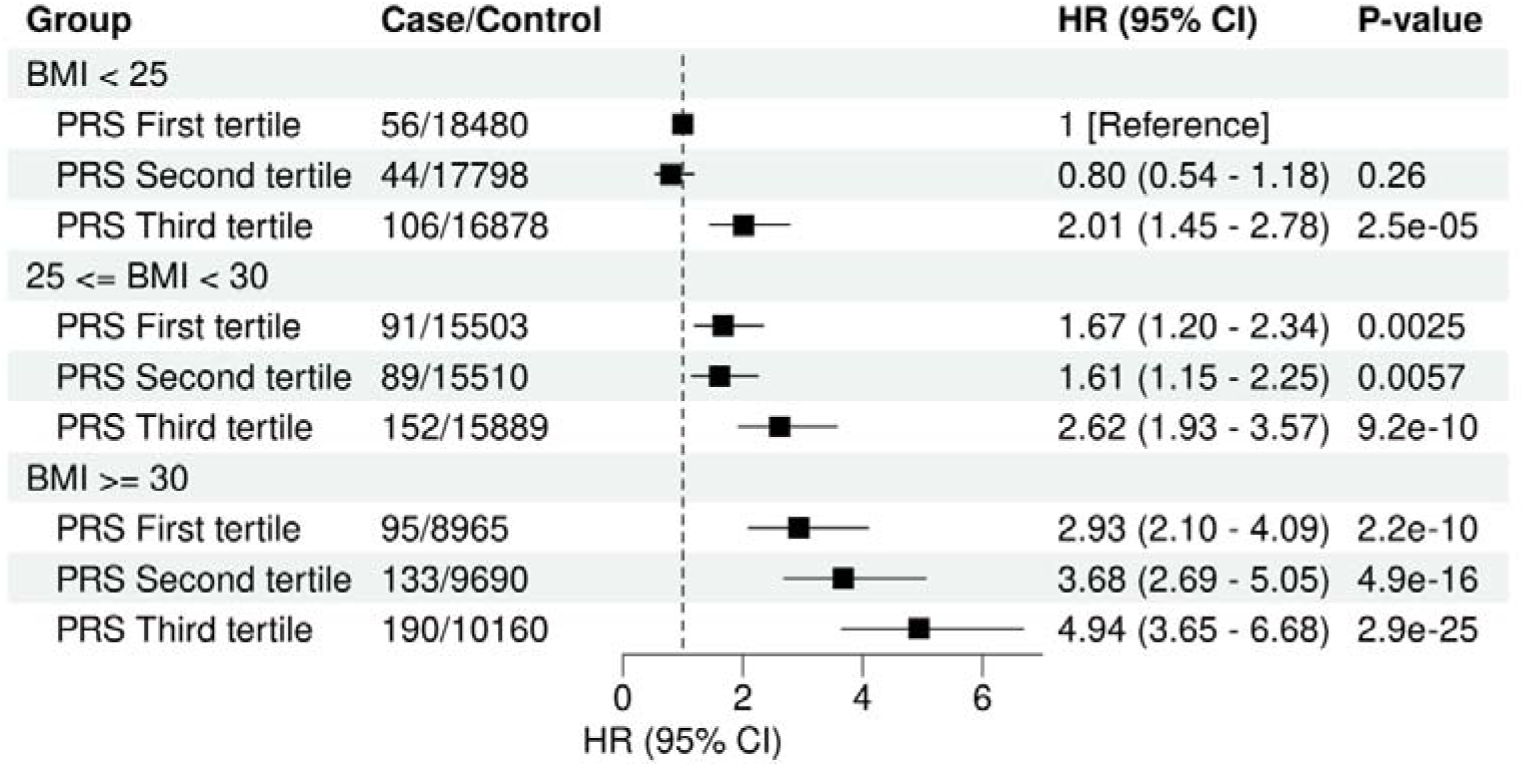
The joint association of genetic risk and BMI with endometrial cancer. Multivariable models were adjusted for age at initial assessment, age at menarche, number of live births, ever taken oral contraceptive pill, PRS values of age at menopause and SHBG and testosterone and the top 10 genetic principal components. The dashed vertical line indicates a Hazard ratio of 1.

## Discussion

In this cohort study of the UK Biobank, our initial focus was on assessing the predictive performance of endometrial cancer PRS and established risk factors. Firstly, we generated a risk distribution for PRS, revealing that individuals in the top 1-10% of the PRS had a risk comparable to that associated with a first-degree family history of endometrial cancer^33,34^. The incorporation of PRS with the epidemiological risk factors in a risk prediction model led to a modest improvement in performance compared to the model that included the non-genetic risk factors alone. Subsequently, we evaluated associations of BMI and PRS with endometrial cancer, finding a joint association with endometrial cancer risk, with obese participants who also had the highest PRS exhibiting the greatest risk. Participants in the top PRS tertile experienced a lower risk if they were not obese. Conversely, individuals in the top PRS tertile had increased endometrial cancer risk compared to other tertiles, even when they had a normal weight. These findings highlight the potential use of endometrial cancer PRS to identify high-risk individuals in the UK Biobank, which is particularly significant given the lack of clinical guidelines for endometrial cancer screening in the general population^35^. Indeed, current screening recommendations are primarily targeted at women with or at risk of Lynch syndrome who account for only ∼3% of endometrial cancer cases.^36,37^

Previous studies have shown that PRS alone provides limited benefit in population screening, individual risk prediction, and population risk stratification.^38,39^ However, disease risk stratification can be improved through the integration of PRS and other risk factors,^40^ as we have also demonstrated. One more appropriate application of PRS is to facilitate personalized cancer screening and management practices. For instance, inclusion of PRS into breast cancer risk estimation may reduce screening and enable more personalised risk management strategies for *CHEK2* and *ATM* pathogenic variant carriers.^41^ PRS has also been shown to add value in distinguishing type 1 from type 2 and monogenic diabetes in adults with a pre-existing diagnosis of diabetes and in selecting the best treatment for different types of diabetes.^42^ As we observed marginal improvement by incorporating endometrial cancer PRS with established risk factors, endometrial cancer PRS may aid in stratifying individuals with Lynch syndrome pathogenic variants.

To the best of our knowledge, this is the first study to quantify the association of BMI with endometrial cancer risk across different genetic risk levels defined by PRS. Our results suggest that, irrespective of PRS, endometrial cancer risk is positively associated with BMI and can be substantially mitigated by weight reduction. An analysis of the Women’s Health Initiative observational study found that women experiencing weight loss of ≥ 5% was linked to a 29% decrease in endometrial cancer risk, and intentional weight loss of ≥ 5% in obese females was associated with 56% lower endometrial cancer risk.^43^ A separate meta-analysis of 13 published studies confirmed a similar association between intentional weight loss and reduced endometrial cancer risk; additionally, the analysis revealed that bariatric surgery was associated with a remarkable 59% reduction in endometrial cancer risk.^44^

The findings that endometrial cancer risk associates with a higher PRS after accounting for established risk factors suggest that endometrial cancer can progress via other mechanisms. Furthermore, although obesity is often defined by BMI, it does not fully represent the distribution of body fat within the human body. In a recent study, it was reported that BMI formed a distinct cluster of fat distribution traits, such as visceral-to-subcutaneous adipose tissue ratio, waist-to-hip ratio without adjustment for BMI, and waist adjusted for BMI. The study further revealed that one unit increase in waist-to-hip ratio or visceral-to-subcutaneous adipose tissue ratio was associated with elevated hazard ratio for endometrial cancer.^45^ Therefore, including other aspects of obesity such as fat distribution and other risk factors not assessed in this study such as duration of hormonal therapy usage, LDL-cholesterol levels, circulating estrogen levels, sedentary behaviours, and smoking (summarized in ^10,11^) may further improve our capability to predict endometrial cancer in the general population.

Our study has several limitations. Firstly, as only a very small proportion of participants had repeated measures or reports for BMI, we could not assess the effects of longitudinal changes of BMI on endometrial cancer risk. Secondly, we only included some known risk factors in our models. Other risk factors not included in this study such as fat distribution and physical activities may modify the joint association of BMI and PRS with endometrial cancer risk. Thirdly, due to the lack of endometrial cancer GWAS in non-European populations, we were unable to assess the ability of risk models to distinguish endometrial cancer in non-European populations.

## Conclusions

The findings of this study demonstrate that an endometrial cancer prediction model incorporating both epidemiological risk factors and PRS has the best performance. The improvement of prediction of endometrial cancer by PRS is anticipated to increase as additional genetic risk variants are identified by larger GWAS. Importantly, BMI and endometrial cancer PRS exert independent effects on the risk of developing endometrial cancer, revealing a substantial increase in risk for obese individuals with high PRS. While underscoring the potential protective impact of weight loss, our findings also indicate that elevated PRS poses an increased risk, even in individuals with a normal weight. These insights emphasise the complex interplay between genetic susceptibility, lifestyle factors, and obesity in endometrial cancer risk, reinforcing the need for personalized and nuanced approaches to screening and preventive interventions.

## Supporting information

eTable 1; eTable 2; eFigure 1

## Data Availability

This research project (Project Application Number 25331) was approved by the UK Biobank in accordance with their established access procedure. UK Biobank data is available to bona fide researchers for health-related research in the public interest. Endometrial cancer GWAS summary statistics used to generate endometrial cancer polygenic risk scores can be downloaded from the GWAS Catalog (Study accession: GCST006464). GWAS summary statistics used to generate polygenic scores for age at natural menopause can be downloaded from the GWAS Catalog (Study accession: GCST90320256). Female-stratified GWAS summary statistics used to generate polygenic scores for SHBG and testosterone can be downloaded from the GWAS Catalog (Study accession: GCST90012107 for SHBG and GCST90012112 for testosterone). Polygenic scores generate in this study will be available at the PGS Catalog upon publication.

https://www.ebi.ac.uk/gwas/studies/GCST006464

https://www.ebi.ac.uk/gwas/studies/GCST90320256

https://www.ebi.ac.uk/gwas/studies/GCST90012107

https://www.ebi.ac.uk/gwas/studies/GCST90012112

## List of abbreviations

PRS: polygenic risk score
PGS: polygenic score
BMI: body mass index
GWAS: genome-wide association study

## Supplementary materials

**eTable 1: Endometrial cancer risk by PRS percentiles**

**eTable 2: Comparison of different predictors to discriminate endometrial cancer cases in the UK Biobank**

**eFigure1: The joint association of genetic risk and BMI with endometrial cancer with additional adjustment for continuous BMI**

## Author Contributions

Drs Wang and O’Mara had full access to all the data in the study and take responsibility for the integrity of the data and the accuracy of the data analysis.

Concept and design: Wang, O’Mara.

Acquisition, analysis, or interpretation of data: All authors.

Drafting of the manuscript: Wang.

Critical revision of the manuscript for important intellectual content: All authors.

Statistical analysis: Wang, O’Mara.

Administrative, technical, or material support: O’Mara.

Supervision: O’Mara.

## Disclaimer

Where authors are identified as personnel of the International Agency for Research on Cancer/World Health Organization, the authors alone are responsible for the views expressed in this article and they do not necessarily represent the decisions, policy or views of the International Agency for Research on Cancer /World Health Organization.

## Reference

1. Bray F, Laversanne M, Sung H, et al. Global cancer statistics 2022: GLOBOCAN estimates of incidence and mortality worldwide for 36 cancers in 185 countries. CA Cancer J Clin. May-Jun 2024;74(3):229–263. doi:10.3322/caac.21834

2. Gu B, Shang X, Yan M, et al. Variations in incidence and mortality rates of endometrial cancer at the global, regional, and national levels, 1990-2019. Gynecol Oncol. May 2021;161(2):573–580. doi:10.1016/j.ygyno.2021.01.036

3. Bafligil C, Thompson DJ, Lophatananon A, et al. Development and evaluation of polygenic risk scores for prediction of endometrial cancer risk in European women. Genet Med. Sep 2022;24(9):1847–1856. doi:10.1016/j.gim.2022.05.014

4. Hüsing A, Dossus L, Ferrari P, et al. An epidemiological model for prediction of endometrial cancer risk in Europe. Eur J Epidemiol. Jan 2016;31(1):51–60. doi:10.1007/s10654-015-0030-9

5. Kachuri L, Graff RE, Smith-Byrne K, et al. Pan-cancer analysis demonstrates that integrating polygenic risk scores with modifiable risk factors improves risk prediction. Nat Commun. Nov 27 2020;11(1):6084. doi:10.1038/s41467-020-19600-4

6. Shi J, Kraft P, Rosner B, et al. Risk prediction models for endometrial cancer: development and validation in an international consortium. J Natl Cancer Inst. Jan 23 2023;doi:10.1093/jnci/djad014

7. Torkamani A, Wineinger NE, Topol EJ. The personal and clinical utility of polygenic risk scores. Nat Rev Genet. Sep 2018;19(9):581–590. doi:10.1038/s41576-018-0018-x

8. Setiawan VW, Yang HP, Pike MC, et al. Type I and II endometrial cancers: have they different risk factors? J Clin Oncol. Jul 10 2013;31(20):2607–18. doi:10.1200/JCO.2012.48.2596

9. Bluher M. Obesity: global epidemiology and pathogenesis. Nat Rev Endocrinol. May 2019;15(5):288–298. doi:10.1038/s41574-019-0176-8

10. Wang X, Glubb DM, O’Mara TA. 10 Years of GWAS discovery in endometrial cancer: Aetiology, function and translation. EBioMedicine. Feb 23 2022;77:103895. doi:10.1016/j.ebiom.2022.103895

11. Raglan O, Kalliala I, Markozannes G, et al. Risk factors for endometrial cancer: An umbrella review of the literature. Int J Cancer. Oct 1 2019;145(7):1719–1730. doi:10.1002/ijc.31961

12. Cheng THT, Thompson DJ, O’Mara TA, et al. Five endometrial cancer risk loci identified through genome-wide association analysis. Nat Genet. Jun 2016;48(6):667–674. doi:10.1038/ng.3562

13. O’Mara TA, Glubb DM, Amant F, et al. Identification of nine new susceptibility loci for endometrial cancer. Nat Commun. Aug 9 2018;9(1):3166. doi:10.1038/s41467-018-05427-7

14. Spurdle AB, Thompson DJ, Ahmed S, et al. Genome-wide association study identifies a common variant associated with risk of endometrial cancer. Nat Genet. May 2011;43(5):451–455. doi:10.1038/ng.812

15. Painter JN, O’Mara TA, Marquart L, et al. Genetic Risk Score Mendelian Randomization Shows that Obesity Measured as Body Mass Index, but not Waist: Hip Ratio, Is Causal for Endometrial Cancer. Cancer Epidem Biomar. Nov 2016;25(11):1503–1510. doi:10.1158/1055-9965.Epi-16-0147

16. Day FR, Thompson DJ, Helgason H, et al. Genomic analyses identify hundreds of variants associated with age at menarche and support a role for puberty timing in cancer risk. Nat Genet. 2017;49(6):834–841. doi:10.1038/ng.3841

17. Ruth KS, Day FR, Hussain J, et al. Genetic insights into biological mechanisms governing human ovarian ageing. Nature. Aug 19 2021;596(7872)doi:10.1038/s41586-021-03779-7

18. D’Urso S, Arumugam P, Weider T, et al. Mendelian randomization analysis of factors related to ovulation and reproductive function and endometrial cancer risk. BMC Med. Nov 1 2022;20(1):419. doi:10.1186/s12916-022-02585-w

19. Collaborative Group on Epidemiological Studies on Endometrial C. Endometrial cancer and oral contraceptives: an individual participant meta-analysis of 27 276 women with endometrial cancer from 36 epidemiological studies. Lancet Oncol. Sep 2015;16(9):1061–1070. doi:10.1016/S1470-2045(15)00212-0

20. Bycroft C, Freeman C, Petkova D, et al. The UK Biobank resource with deep phenotyping and genomic data. Nature. Oct 2018;562(7726):203–209. doi:10.1038/s41586-018-0579-z

21. Kho PF, Mortlock S, Rogers PAW, et al. Genetic analyses of gynecological disease identify genetic relationships between uterine fibroids and endometrial cancer, and a novel endometrial cancer genetic risk region at the WNT4 1p36.12 locus. Hum Genet. Sep 2021;140(9):1353–1365. doi:10.1007/s00439-021-02312-0

22. Lloyd-Jones LR, Zeng J, Sidorenko J, et al. Improved polygenic prediction by Bayesian multiple regression on summary statistics. Nat Commun. Nov 8 2019;10(1):5086. doi:10.1038/s41467-019-12653-0

23. Zeng J, de Vlaming R, Wu Y, et al. Signatures of negative selection in the genetic architecture of human complex traits. Nat Genet. May 2018;50(5):746–753. doi:10.1038/s41588-018-0101-4

24. Choi SW, Mak TS, O’Reilly PF. Tutorial: a guide to performing polygenic risk score analyses. Nat Protoc. Sep 2020;15(9):2759–2772. doi:10.1038/s41596-020-0353-1

25. Purcell S, Neale B, Todd-Brown K, et al. PLINK: a tool set for whole-genome association and population-based linkage analyses. Am J Hum Genet. Sep 2007;81(3):559–75. doi:10.1086/519795

26. Wang X, Kho PF, Ramachandran D, et al. Multi-trait GWAS identifies a novel endometrial cancer risk locus that associates with testosterone levels. iScience. 2023;26:106590. doi:10.1016/j.isci.2023.106590

27. Ruth KS, Day FR, Tyrrell J, et al. Using human genetics to understand the disease impacts of testosterone in men and women. Nature Medicine. 2020;26(2):252–258. doi:10.1038/s41591-020-0751-5

28. Yang J, Benyamin B, McEvoy BP, et al. Common SNPs explain a large proportion of the heritability for human height. Nat Genet. Jul 2010;42(7):565–9. doi:10.1038/ng.608

29. Yang J, Lee SH, Goddard ME, Visscher PM. GCTA: a tool for genome-wide complex trait analysis. Am J Hum Genet. Jan 7 2011;88(1):76–82. doi:10.1016/j.ajhg.2010.11.011

30. Lee SH, Goddard ME, Wray NR, Visscher PM. A better coefficient of determination for genetic profile analysis. Genet Epidemiol. Apr 2012;36(3):214–24. doi:10.1002/gepi.21614

31. Kundu S, Aulchenko YS, van Duijn CM, Janssens AC. PredictABEL: an R package for the assessment of risk prediction models. Eur J Epidemiol. Apr 2011;26(4):261–4. doi:10.1007/s10654-011-9567-4

32. Robin X, Turck N, Hainard A, et al. pROC: an open-source package for R and S+ to analyze and compare ROC curves. BMC Bioinformatics. Mar 17 2011;12:77. doi:10.1186/1471-2105-12-77

33. Win AK, Reece JC, Ryan S. Family history and risk of endometrial cancer: a systematic review and meta-analysis. Obstet Gynecol. Jan 2015;125(1):89–98. doi:10.1097/AOG.0000000000000563

34. Johnatty SE, Tan YY, Buchanan DD, et al. Family history of cancer predicts endometrial cancer risk independently of Lynch Syndrome: Implications for genetic counselling. Gynecol Oncol. Nov 2017;147(2):381–387. doi:10.1016/j.ygyno.2017.08.011

35. Crosbie EJ, Kitson SJ, McAlpine JN, Mukhopadhyay A, Powell ME, Singh N. Endometrial cancer. Lancet. Apr 9 2022;399(10333):1412–1428. doi:10.1016/S0140-6736(22)00323-3

36. Ryan NAJ, Glaire MA, Blake D, Cabrera-Dandy M, Evans DG, Crosbie EJ. The proportion of endometrial cancers associated with Lynch syndrome: a systematic review of the literature and meta-analysis. Genet Med. Oct 2019;21(10):2167–2180. doi:10.1038/s41436-019-0536-8

37. Ryan NAJ, McMahon R, Tobi S, et al. The proportion of endometrial tumours associated with Lynch syndrome (PETALS): A prospective cross-sectional study. PLoS Med. Sep 2020;17(9):e1003263. doi:10.1371/journal.pmed.1003263

38. Hingorani AD, Gratton J, Finan C, et al. Performance of polygenic risk scores in screening, prediction, and risk stratification: secondary analysis of data in the Polygenic Score Catalog. BMJ Med. 2023;2(1):e000554. doi:10.1136/bmjmed-2023-000554

39. Huntley C, Torr B, Sud A, et al. Utility of polygenic risk scores in UK cancer screening: a modelling analysis. Lancet Oncol. May 10 2023;24(6):658–668. doi:10.1016/S1470-2045(23)00156-0

40. Lee A, Mavaddat N, Wilcox AN, et al. BOADICEA: a comprehensive breast cancer risk prediction model incorporating genetic and nongenetic risk factors. Genet Med. Aug 2019;21(8):1708–1718. doi:10.1038/s41436-018-0406-9

41. Gao C, Polley EC, Hart SN, et al. Risk of Breast Cancer Among Carriers of Pathogenic Variants in Breast Cancer Predisposition Genes Varies by Polygenic Risk Score. J Clin Oncol. Aug 10 2021;39(23):2564–2573. doi:10.1200/JCO.20.01992

42. Luckett AM, Weedon MN, Hawkes G, Leslie RD, Oram RA, Grant SFA. Utility of genetic risk scores in type 1 diabetes. Diabetologia. Sep 2023;66(9):1589–1600. doi:10.1007/s00125-023-05955-y

43. Luo J, Chlebowski RT, Hendryx M, et al. Intentional Weight Loss and Endometrial Cancer Risk. J Clin Oncol. Apr 10 2017;35(11):1189–1193. doi:10.1200/JCO.2016.70.5822

44. Zhang X, Rhoades J, Caan BJ, et al. Intentional weight loss, weight cycling, and endometrial cancer risk: a systematic review and meta-analysis. Int J Gynecol Cancer. Nov 2019;29(9):1361–1371. doi:10.1136/ijgc-2019-000728

45. Rask-Andersen M, Ivansson E, Hoglund J, Ek WE, Karlsson T, Johansson A. Adiposity and sex-specific cancer risk. Cancer Cell. Jun 12 2023;41(6):1186–1197 e4. doi:10.1016/j.ccell.2023.05.010

