## Supplementary material for "Risk Stratification for Endometrial Cancer Reveals Independent Contributions of Polygenic Risk and Body Mass Index": eTable 1; eTable 2; eFigure 1

**Supplementary materials**

**eTable 1 Endometrial cancer risk by PRS percentiles**

**eTable 2 Comparison of different predictors to discriminate endometrial cancer cases in the UK Biobank**

**eFigure 1** **The joint association of genetic risk and BMI with endometrial cancer with additional adjustment for continuous BMI**

**eTable 1 Endometrial cancer risk by PRS percentiles.**

| Percentile | OR | 95% CI | P-value | count of cases | count of non-cases |
| --- | --- | --- | --- | --- | --- |
| 0-10% | 0.64 | 0.47 - 0.89 | 7.38E-03 | 49 | 13,382 |
| 10-20% | 1.04 | 0.79 - 1.37 | 0.78 | 79 | 13,351 |
| 20-30% | 1.08 | 0.82 - 1.41 | 0.58 | 82 | 13,348 |
| 30-40% | 1.16 | 0.89 - 1.51 | 0.27 | 88 | 13,343 |
| 40-60% (Reference) | 1 | NA | 0 | 152 | 26,708 |
| 60-70% | 1.60 | 1.26 - 2.03 | 1.28E-04 | 121 | 13,310 |
| 70-80% | 1.65 | 1.30 - 2.09 | 3.54E-05 | 125 | 13,305 |
| 80-90% | 1.80 | 1.42 - 2.27 | 7.49E-07 | 136 | 13,294 |
| 90-100% | 1.98 | 1.58 - 2.49 | 3.03E-09 | 127 | 11,960 |
| 99-100% | 3.06 | 1.97 - 4.76 | 7.10E-07 | 23 | 1,321 |

Abbreviations - OR: Odds ratio; CI: Confidence interval.

**eTable 2 Comparison of different predictors to discriminate endometrial cancer cases in the UK Biobank.**

| Prediction model^a^ | AUC | 95% CI | Specificity | Sensitivity |
| --- | --- | --- | --- | --- |
| Endometrial cancer PRS | 0.671 | 0.654 - 0.687 | 0.588 | 0.660 |
| BMI (continuous) | 0.713 | 0.696 - 0.729 | 0.676 | 0.639 |
| Number of live births (count) | 0.648 | 0.632 - 0.664 | 0.601 | 0.605 |
| Age of menarche | 0.655 | 0.639 - 0.670 | 0.607 | 0.618 |
| Age of menopause PGS | 0.648 | 0.632 - 0.664 | 0.589 | 0.629 |
| Oral contraceptive use (never/ever) | 0.652 | 0.637 - 0.668 | 0.589 | 0.628 |
| SHBG level PGS | 0.659 | 0.644 - 0.675 | 0.582 | 0.650 |
| Testosterone level PGS | 0.652 | 0.636 - 0.668 | 0.601 | 0.618 |
| Epidemiological model^b^ | 0.728 | 0.712 - 0.744 | 0.647 | 0.683 |
| Integrative model^c^ | 0.739 | 0.723 - 0.754 | 0.716 | 0.633 |

Abbreviations - AUC: area under the receiver operator curve; CI: confidence interval; PRS: polygenic risk score; BMI: body mass index; PGS: polygenic score; SHBG: sex hormone binding globulin.

^a^All models were adjusted for age at initial visit, assessment centre and the first 10 principal components.

^b^Epidemiological model included BMI, age of menarche, oral contraceptive use, number of live births, age of menopause PGS, SHBG level PGS and testosterone level PGS.

^c^Integrative model included the epidemiological model plus the endometrial cancer PRS.

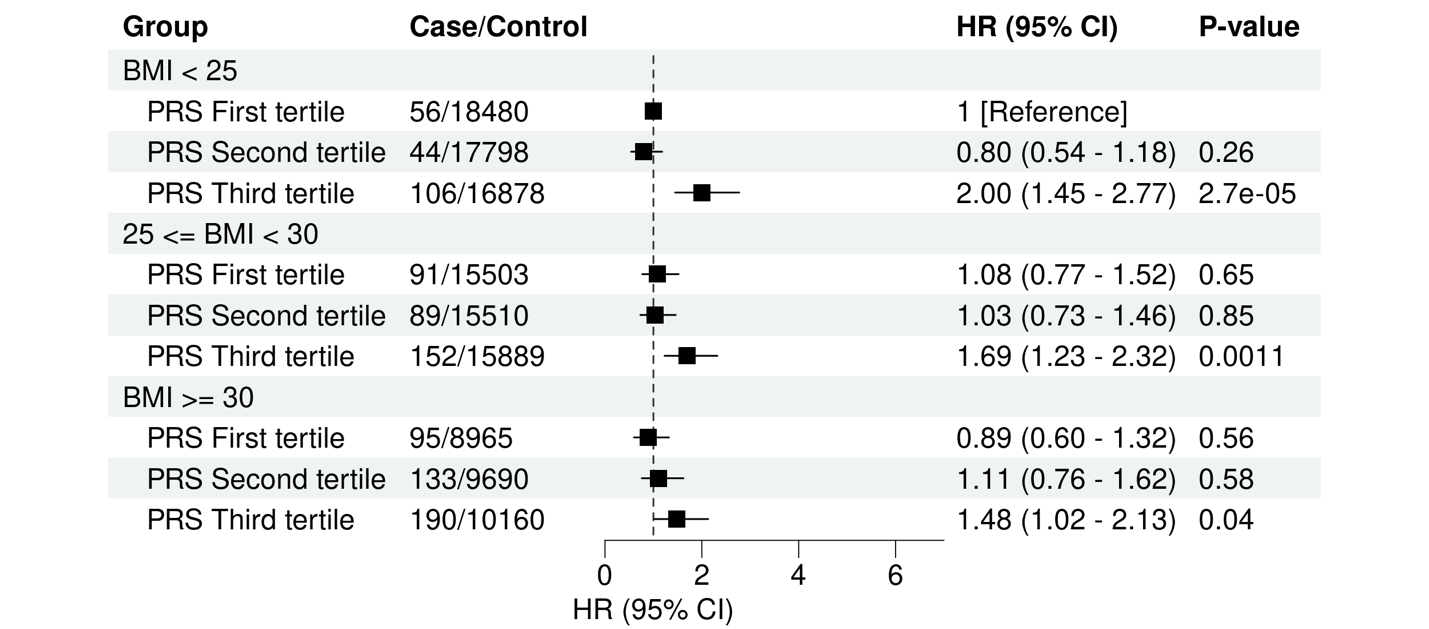

**eFigure1** **The joint association of genetic risk and BMI with endometrial cancer with additional adjustment for continuous BM.**
